# Ethical Considerations When Creating Evidence from Real World Digital Health Data

**DOI:** 10.1101/2020.01.16.20017806

**Authors:** C. Nebeker, Victoria Leavy, Eva Roitmann, Steven Steinhubl

**Author notes:** **Correspondence:** Camille Nebeker, EdD, MS, Associate Professor, Department of Family Medicine & Public Health, School of Medicine, UC San Diego, La Jolla, CA 92093-0811, 01-858-534-7786.

## Abstract

**Background:** Personal health data (PHD) are collected using digital self-tracking technologies and present opportunities to increase self-knowledge and, also biometric surveillance. PHD become “big” data and are used in health-related research studies. We surveyed consumers regarding expectations regarding consent and sharing of PHD for biomedical research.

**Methods:** Data sharing preferences were assessed via an 11-item survey. The survey link was emailed to 89539 English-speaking Withings product users. Responses were accepted for 5 weeks.

Descriptive statistics were calculated using Excel and qualitative data were analyzed to provide additional context.

**Results:** Nearly 1640 people or 5.7% of invitees responded representing 62 countries with 80% identifying as Caucasian, 75% male with 78% being college educated. The majority were agreeable to having their data shared with researchers to advance knowledge and improve health care.

Participants responding to open ended items (N=247) appeared unaware that the company had access to their personal health data.

**Conclusions:** While the majority of respondents were in favor of data sharing, individuals expressed concerns about the ability to de-identify data and associated risks of re-identification as well as an interest in having some control over the use of “their” data. Given consumer misconception about data ownership, access and use, efforts to increase transparency when interacting with individual digital health data must be prioritized. Moreover, the basic ethical principle of “respect for persons” demonstrated via the informed consent process will be critical in advancing the adoption of digital technologies that create real-world evidence and advance opportunities for N-of-1 self-study.

## Introduction

A longstanding tradition and ethical mandate of biomedical research is the act of obtaining informed consent. Informed consent involves providing information about the research prospectively with a goal of arming a person who is considering study participation with the relevant details needed to make an informed decision to volunteer. There are times when obtaining informed consent in advance of study enrollment is not feasible (e.g., emergency medicine) and, as such, there are regulations that permit the waiver of informed consent, which can be considered contradictory and ethically questionable (US Food and Drug Administration, 2016; Klein, et al. 2018). There is also a regulatory exemption category in that permits the use of existing data for secondary analyses when the person cannot be identified directly or through identifiers (US Department of Health and Human Services). These rules pertain to research that is conducted by individuals or organizations bound by the United States (US) Code of Federal Regulations that have a Federal Wide Assurance of Compliance to carry out Department of Health and Human Services supported research. Not all entities conducting health research are bound by these rules and, as technology companies amass enormous quantities of consumer information we are beginning to see the use of consumer data in an unregulated health research environment. In unregulated research, adherence to federal regulations, including the use of an institutional review board (IRB) may not be required nor sought out.

In 2016, the results of an observational study were published reporting on holiday weight gain using data obtained from users of a wireless scale (WS50, Withings). The study goal was to observe weight change associated with holidays in the US, Japan and Germany using de-identified data collected from users of the scale (Helander, et al. 2016). Curious about consumer perspectives around sharing health-related data for biomedical research, we surveyed users of a variety of Withings consumer health devices to learn about their data sharing preferences and expectations.

## Methods

Participant had at least one Withings device with recent Health Mate app activity. A short survey soliciting data sharing preferences was developed by the co-authors. A Likert-scale was used where 1= Strongly Disagree, 2= Disagree, 3= Neither Agree nor Disagree, 4= Agree, and 5= Strongly Agree, with six questions focused on sharing data with researchers and four on how their health care provider may benefit from data access (see Appendix A).

Following each question set, participants could comment to qualify their thoughts about sharing data for research and health care purposes. Descriptive statistics were calculated using Excel and qualitative data were analyzed to provide additional context. The study was verified as exempt (see 45 CFR 46.101) by the [blinded for peer review] IRB and launched April 2018. Withings personnel disseminated the survey link via email to 89539 English-speaking users of whom 28601 opened the e-mail. Responses were accepted for 5 weeks.

## Results

Nearly 1640 people, or 5.7% of those who received and opened the mail, responded to the survey representing 62 countries with 80% identifying as Caucasian, 75% male and 78% having a bachelor degree or graduate education. Results reveal that the majority are agreeable to having their data shared with researchers, if it could advance knowledge and improve health care (N=1639, mean 4.27, SD 1.1). There was slightly more interest in sharing of de-identified data (N=1639, mean 4.38, SD 1.0) and, if that were to occur, participants expressed a strong interest in knowing what was learned by receiving a study summary (N=1631, mean 4.44, SD 1.0) and a copy of results once published (N=1636, mean 4.23, SD 1.15). When asked if they carefully read service agreements when deciding to use an app or device, the majority indicated disagreement (N=1636, mean 2.8, SD 1.3). Slightly more participants expressed an interest in suggesting research questions that could be answered using the company apps and devices (N=1633, mean 3.27, SD1.3). Participants were agreeable to having their care provider access health data obtained via commercial devices, if it led to better care (N=1636, mean 4.09, SD 1.1) and would approve of sharing data with providers (N=1635, mean 4.09, SD 1.1) but, were not certain their health care providers would be interested in having data access (N=1633, mean 3.80, SD 1.17). Lastly, the participants were mixed on whether clinician access would influence their decision to select a hospital or health care provider (N=1637, mean 3.77, SD 1.3) (See Table 1).

**Table 1:** Descriptive statistics of 10-item survey.

|  | N | Mean |  | Std. Deviation |
| --- | --- | --- | --- | --- |
|  | Statistic | Statistic | Std. Error | Statistic |
| q1_for_health | 1639 | 4.27 | .027 | 1.110 |
| q2_when_deidentified | 1639 | 4.38 | .027 | 1.097 |
| q3_summary_of_findings | 1631 | 4.44 | .026 | 1.041 |
| q4_medical_publication | 1636 | 4.23 | .029 | 1.154 |
| q5_read_terms | 1636 | 2.83 | .033 | 1.337 |
| q6_suggesting_research | 1633 | 3.27 | .033 | 1.330 |
| q8_better_care | 1636 | 4.09 | .028 | 1.139 |
| q9_ability_to_share | 1635 | 4.09 | .030 | 1.199 |
| q10_care_provider_interested | 1633 | 3.80 | .029 | 1.177 |
| q11_chose_provider_if_use_data | 1637 | 3.77 | .032 | 1.304 |

Approximately 15% (n=247) of participants provided comments. While a comprehensive qualitative analysis is beyond the scope of this perspectives essay, examples are provided for context. People indicating strong agreement with sharing were generally supportive of the use of data as noted by these examples: “I fully support using collected health data to further scientific research to improve health.” (P 668) Yet, supporters also had contingencies about sharing, which focused primarily on compensation and regulations. For example, “If an organization is making money from my data, I should be fairly compensated for my data. If freely given to researchers, then I don’t require compensation.” (P 394) and “It would be a good thing, if users would benefit either by getting discounts or paid in cash if their data is used.” (P 513) Regulatory concerns were mostly privacy related: “I would strongly agree if the research is based in the EU, if it is in USA or other countries with less regulation of privacy of data I would not allow the sharing of health data at all.” (P 282)

Examples of those unwilling to share include: “I do not want my private health care details shared with anyone for any reason.” (P 1442) and “I do not want my data shared whatsoever!!!!!!!!!!” (P 1332). Similar to supporters, privacy concerns were expressed, “You do not have permission to share my data. Comply with EU privacy rules. There is no such thing as de-identifying data, it can all too easily be de-anonymized.” (P 146) And, a few referenced Facebook “Until I am convinced that there are adequate standards and laws in place to prevent Facebook-type abuses I will be very reluctant to share any identifiable information. Privacy must be preserved. Transparency must be total.” (P 1205)

## Discussion and Conclusion

This survey was launched shortly after the Facebook/Cambridge Analytica data breach occurred, which may have influenced participant responses. While the majority of respondents were in favor of data sharing, individuals expressed concerns about the ability to de-identify data and associated risks of reidentification as well as an interest in having some control over the use of “their” data. While it may not be surprising, the comments indicate that some consumers may not be aware that their data is obtained and stored by the entity providing the app or device. At least for some people, this lack of knowledge may lead to less trust and potentially compromise consumer willingness to use digital health products.

Moving forward, when prospective consent to partake in research is not obtained because there are no mandates, or the permissions are buried in the terms of service (ToS) agreements, there are consequences to consider if privacy expectations are compromised or a data breach occurs. In a study by Merz et al. (2018), 30% of people, if given the right to consent, would opt out (Baker, Merz, 2018). The Facebook Emotional Contagion study, OK Cupid and now Cambridge Analytica research studies have led to a backlash from consumers that has the potential to jeopardize important future health research if consumer trust continues to deteriorate. Efforts to increase transparency when interacting with consumers combined with technology education is a starting point to advancing a big data research agenda and enhance public trust of science. In that End User Licensing Agreements and ToS are the primary method of obtaining legal “acceptance” (not to be confused with informed consent) from a user within the commercial realm, striving to strike a balance between the legal (i.e. reducing exposure to liability and ensuring business interests are achieved) and ethical (i.e. ensuring consumer comprehension and voluntariness) interests may be a first step towards greater transparency.

Author ER was employed by the company Withings. The remaining authors declare that the research was conducted in the absence of any commercial or financial relationships that could be construed as a potential conflict of interest.

## Data Availability

Quantitative data and qualitative data we be provided after redaction of identifiable information and are available through the Scripps Translational Science Institute.

## Appendix A

### Survey Items

Survey questions to assess consumer willingness to share their personal health data with researchers and health care providers.

|  |
| --- |
| **Researcher Access** |
| 1. If my health data can help advance health understanding and care improvement, I am generally supportive of sharing it. |
| 2. Knowing that my data is de-identified and there is no way to link it with my identity, I am more likely to share my health data with researchers. |
| 3. When my de-identified data is used to advance health understanding and care improvement, I'd like to receive a summary of the study findings. |
| 4. When my de-identified data is used to advance health understanding and care improvement, I'd like to receive a copy of the results once they are published in a medical research journal. |
| 5. In general, I carefully read the terms of service agreements that all consumers must accept in order to use a device or app. |
| 6. I am interested in suggesting health-related research questions that could be answered by analyzing health data from the Nokia Health Mate app and devices. |
| **Clinician Access** |
| 7. I believe that my care provider(s) would be able to provide me with better health care if they could access data from my health devices (such as weight, heart rate etc.). |
| 8. I would like to be able to share the data collected by my health devices with my care provider(s). |
| 9. To provide me with better care, I believe that my care provider(s) would be interested in having access to data from my health devices. |
| 10. I would be more likely to select a care provider or hospital for my care if they offered to receive and interpret the data collected by my health devices. |

